# Parameters leading to cerebrospinal fluid diversion in low-grade subarachnoid hemorrhage – a Neurobase study

**DOI:** 10.1101/2025.11.26.25341125

**Authors:** Marie Lejeune, Benjamin G. Chousterman, Samy Figueiredo, Jonas Pochard, Stéphanie Sigaut, Bertrand Mathon, Matthieu Landart, Adam Biard, Léa Meyer, Sara Van Phan, Vincent Degos, Alice Jacquens, Rémy Bernard

## Abstract

**Background and Purpose:** Patients admitted for subarachnoid hemorrhage (SAH) may require cerebrospinal fluid (CSF) diversion. The rate and indications for CSF diversion are highly variable. This study aimed to identify factors associated with CSF diversion in low-grade SAH in clinical practice.

**Methods:** We used a prospectively collected multicentric database (Neurobase®) cohort of patients with severe acute cerebral injuries, in three intensive care units (ICU) in France between March 2022 and June 2024. We included patients with aneurysmal or *sine materia* SAH with WFNS grades 1 to 3. Clinical and radiological variables were analyzed in patients with a CSF diversion during ICU stay. Hydrocephalus, cerebral hemorrhage and edema were quantified on admission CT-scans.

**Results:** Our cohort included 219 patients, mainly women (61%), on average 53 years old, mostly WFNS grade 1 (68%). Shunts were placed in 45% of patients, of which 92% were external ventricular drains, and 75% were placed in the first 24 hours. The predictors of CSF diversion in the first 24 hours in multivariate analysis were the delay between symptoms and first CT-scan (adjusted OR [aOR], 0.67 [95% CI, 0.45-0.99]), a GCS in the first 24 hours < 15 (aOR, 3.17 [95% CI, 1.42-7.10]), the ventricular Hijdra score (aOR, 1.48 [95% CI, 1.19-1.85]), temporal horn dilation (aOR, 1.89 [95% CI, 1.25-2.87]), and a Subarachnoid hemorrhage Early Brain Edema Score (SEBES) of 2 (aOR, 4.83 [95% CI, 1.57-14.87]). These predictors were also associated with CSF diversion at any time of ICU stay. We tested the centers and found a statistical significance (p < 0.001).

**Conclusion:** The decision to place a CSF shunt depends not only on hydrocephalus, altered mental status, and intraventricular hemorrhage, but also on cerebral edema and the delay between the first symptoms and CT-scan. The variability of practice in the included centers highlights a gray zone in the indication of CSF diversion.

## INTRODUCTION

Subarachnoid hemorrhage (SAH) is a subtype of hemorrhagic stroke, most frequently due to a ruptured aneurysm of an intracranial artery^1^. Acute hydrocephalus is one of the most common complications of aneurysmal SAH. Current European and US guidelines recommend cerebrospinal fluid (CSF) diversion by placement of an external ventricular drain (EVD) or lumbar drainage in case of acute hydrocephalus associated with an altered mental status (EU : recommendation class IV, evidence level C ; US : recommendation class 1, evidence level B-NR [moderate-quality evidence from non-randomized studies]) or intraventricular hemorrhage (EU : good clinical practice, low level of evidence)^2,3^. However, the incidence of acute hydrocephalus in aneurysmal SAH varies widely in published studies from 12% to 87%, as does the incidence of EVD placement from 25% to 66% in low-grade aneurysmal SAH (defined as a World Federation of Neurological Surgeons (WFNS) grade or Hunter-Hess score ≤ 3)^4,5^. This may be due to the lack of a standardized definition of acute hydrocephalus. Although many radiological criteria have been described (bicaudate index, third ventricle dilation, temporal horn dilation), acute hydrocephalus is often diagnosed in routine clinical practice by visual and subjective qualification on cerebral imaging, which may be biased by physicians’ experience^6^. The decision to place a CSF shunt may therefore vary according to local protocols and experiences.

The benefits of a CSF diversion in patients with low-grade SAH should be balanced against the common and potentially life-threatening risks associated with EVD placement, such as hemorrhagic complications including aneurysm rebleeding from rapidly alleviating intracranial pressure, infectious complications including meningitis or ventriculitis, and revision due to catheter misplacement or obstruction. These complications have been reported to occur in up to 30% of patients^7^.

We hypothesized that CSF diversion is decided not only on hydrocephalus and altered mental status but may also depend on other factors that have yet to be identified. The main objective of this study was to determine predictive factors for CSF diversion in the first 24 hours from intensive care unit (ICU) admission, and secondarily during the entire ICU stay, in patients with low-grade non-traumatic SAH.

## METHODS

### Study Population

We performed a descriptive observational retrospective study with data prospectively collected from a multicentric SAH cohort of patients admitted to three specialized neurosurgical ICUs in France, from March 1, 2022, to June 30, 2024. All adult patients admitted with aneurysmal or *sine materia* SAH with a WFNS grade of 1, 2 or 3, were included in this study. Exclusion criteria were the presence of another cause of SAH (such as, but not limited to, arteriovenous malformation, traumatic SAH), early rebleeding before intervention leading to a Glasgow Coma Scale (GCS) < 13, preexisting hydrocephalus or CSF shunt, and pregnant or breastfeeding women.

### Ethics

The data was collected from patients’ medical files and from a database named Neurobase®, registered under the number CER-2021-77, which was examined and approved in 2021 by the ethics committee of Assistance Publique-Hôpitaux de Paris (Institutional Review Board – IRB 00006477 – de l’HUPNVS, Université Paris 7, AP-HP) in accordance with the Declaration of Helsinki. STROBE guidelines were followed in the writing of this article.

### Data on CSF Diversion Characteristics

The data collected for CSF diversion included the type of shunt (EVD, lumbar drainage, and lumbar puncture), the delay between ICU admission and diversion, the presence of complications related to diversion, and the necessity of a permanent ventriculo-peritoneal or ventriculo-atrial shunt.

### Admission Head CT-scan

Acute hydrocephalus was defined by three radiological variables on the first head CT-scan: age-relative bicaudate index, temporal horn dilation (collected as a semi-quantitative variable), and the width of the third ventricle in millimeters^6,8,9^. Intracerebral hemorrhage was measured with the modified Graeb score and the ventricular and subarachnoid Hijdra scores^10,11^. Cerebral edema was evaluated with the Subarachnoid hemorrhage Early Brain Edema Score (SEBES)^12^.

### Guideline-Based Indication of CSF Diversion

Formal indication of CSF diversion was defined as having a GCS lower than 15 associated with moderate or severe temporal horn dilation, and no formal indication of CSF diversion was defined as having a GCS of 15 associated with absent or minimal temporal horn dilation. Patients that did not fit in either category were defined as being in a gray area of indication.

### General Data

Patients’ general characteristics were collected, including age, sex, modified Fisher score, lowest GCS in the first 24 hours, and the delay between the first onset of symptoms and the first head CT-scan. The worst pulsatility index (PI) on transcranial Doppler (TCD) was also collected as a clinical measure of intracranial hypertension.

Patients were followed up to one year after discharge, and good functional outcome was defined by a modified Rankin scale (mRS) of 0 to 3 or a Glasgow Outcome Scale (GOS) of 4 to 5.

### Statistical Analysis

Statistical analysis was performed using R (version 4.3.2).

Depending on the type of variable and the normality of its distribution, the results are presented as a mean with standard deviation (SD), a median with interquartile interval, or number of subjects associated with its proportion. Bivariate comparisons of the predictive factors of CSF diversion required exact Fisher tests for discrete variables and Student or Wilcoxon tests for continuous variables depending on their distribution. Missing data was described and not analyzed.

To identify the predictive factors of CSF diversion, a univariate analysis followed by multivariate logistic regression models were performed. A first predictive model for CSF diversion within the first 24 hours of admission and another predictive model for CSF diversion during the entire ICU stay were developed as the primary and secondary objectives of the study.

The log-linearity of continuous variables was verified, and these variables were modified as discrete variables if needed before their integration into the multivariate logistic regression models. We therefore modified the GCS variable into a dichotomous one (15 or less than 15), the ventricular Hijdra score into a quantitative variable truncated at 6, and the SEBES score into a qualitative variable. The variables that were included in the multivariate logistic regression models were selected based on a strong *a priori* (age and *sine materia* SAH) or a p-value < 0.20 in the univariate analysis, and the radiological variables that were most predictive of hydrocephalus, intracerebral hemorrhage, and cerebral edema. The receiver operating characteristic (ROC) curves were established for these variables, and the area under the curve (AUC) was calculated. Since multiple or colinear variables described hydrocephalus or intracerebral hemorrhage, the ones with the best discriminating value on the AUC in the univariate analysis were chosen. The quality of the final model was evaluated by the AUC representing its discriminatory performance and calibration curves. We added the center variable to the final model as a fixed effect variable to test for possible heterogeneity between centers.

The reliability of the modified Fisher score and of the appreciation of temporal horn dilation was tested by measuring the agreement between 2 physicians (including a neurosurgeon) using weighted Cohen’s Kappa test.

All tests were bilateral, and likelihood-ratio tests were performed in multivariate analysis to determine the degree of significance of the predictive factors. The threshold for the p-value was set at 0.05.

## RESULTS

### Patients’ Characteristics

Between March 2022 and June 2024, 263 patients with low-grade aneurysmal or *sine materia* SAH were included in the database. Of those patients, 44 were excluded from our analysis (mostly for early rebleeding with GCS < 13 or unavailable first head CT-scan), which allowed for the analysis of 219 patients. The flow chart is described in Figure 1.

**Figure 1:**
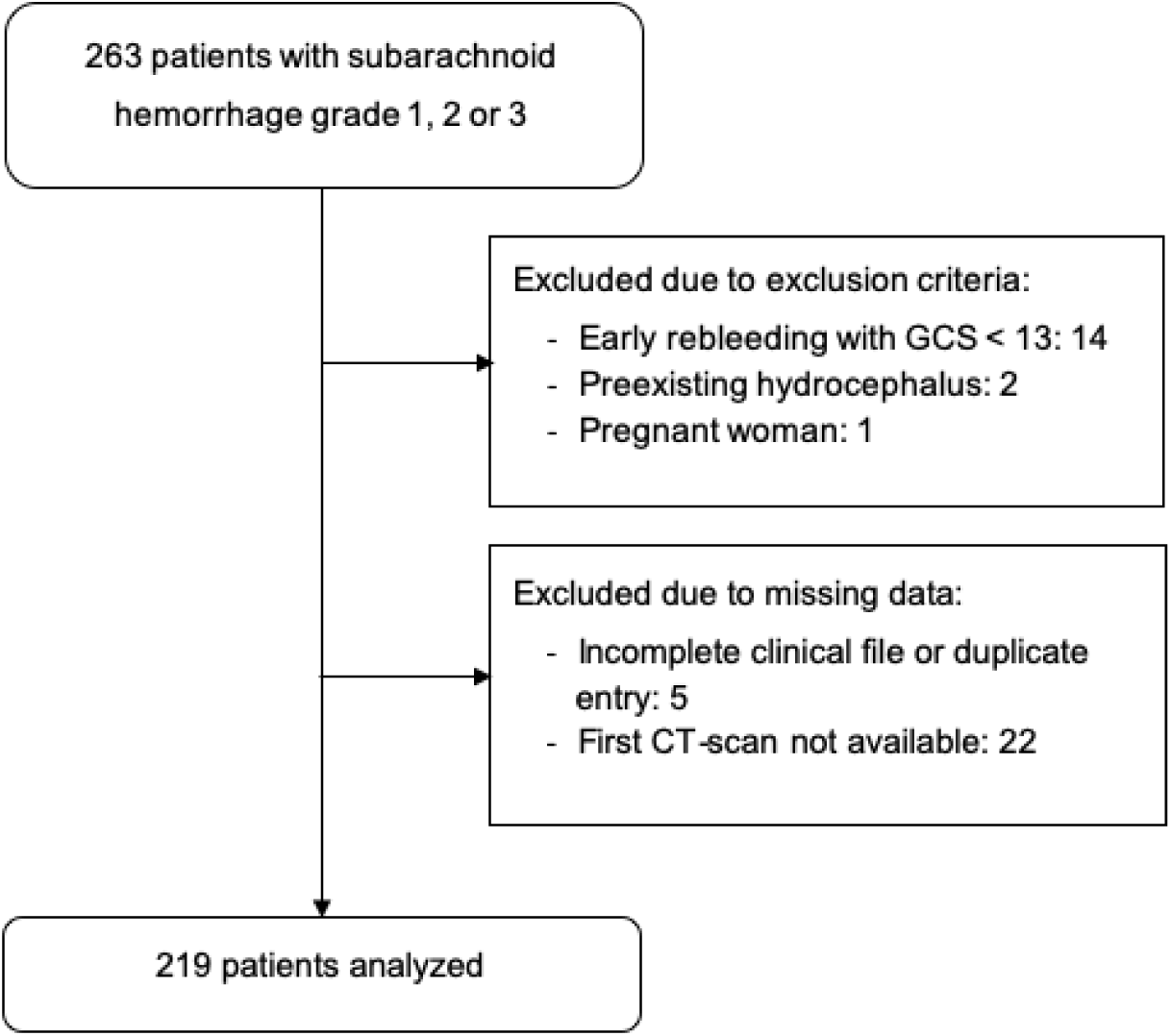
Flow Chart.

In this cohort, 60.7% of patients were female with a mean age of 52.9 years old, and 68% had a WFNS grade 1 (Table 1). A CSF shunt was placed at any time during the ICU stay in 99 patients (45.2%), with mainly EVDs (93.1%). The description of CSF diversion characteristics in this cohort is shown in Table 2. CSF diversion occurred before aneurysmal treatment in 72.7% of patients, and most shunts were placed early in the first 48 hours following ICU admission (91.9%), with 74.7% in the first 24 hours (Figure S1).

**Table 1:** Patient Characteristics.

| Characteristic |  | N = 219 |
| --- | --- | --- |
| Center, n (%) | 1 | 42 (19.2) |
|  | 2 | 79 (36.1) |
|  | 3 | 98 (44.7) |
| Age (years), mean (SD*) |  | 52.8 (13.0) |
| Sex, n (%) | Female | 133 (60.7) |
| Hypertension, n (%) |  | 73 (34.0) |
| Diabetes, n (%) |  | 15 (6.9) |
| WFNS† grade, n (%) | 1 | 149 (68.0) |
|  | 2 | 62 (28.3) |
|  | 3 | 8 (3.7) |
| Lowest GCS‡ in first 24h, n (%) | < 13 | 19 (8.4) |
| Type of aneurysmal treatment, n (%) | Endovascular | 155 (71.8) |
|  | Surgical | 22 (10.2) |
|  | *Sine materia* | 39 (18.1) |
| Duration of stay (days), mean (SD) |  | 14.3 (9.2) |
| Death, n (%) |  | 7 (3.1) |
| Functional outcome, n (%) | Favorable | 200 (91.3) |

\*SD: Standard Deviation; †WFNS: World Federation of Neurological Surgeons; ‡GCS: Glasgow Coma Score

**Table 2:** Details on CSF Diversion.

| CSF* diversion characteristic |  | n = 99 |
| --- | --- | --- |
| Delay between admission and | < 24 hours | 74 (74.7) |
| CSF diversion, n (%) | < 48 hours | 91 (91.9) |
| CSF diversion before aneurysmal treatment, n (%) |  | 72 (72.7) |
| Type of CSF shunt, n (%) | EVD† | 92 (92.9) |
|  | EVD associated with lumbar catheter drainage | 14 (14.1) |
|  | Lumbar catheter drainage without EVD | 1 (1.0) |
|  | Lumbar puncture without catheter | 6 (6.1) |
| CSF diversion complications, n (%) | > 1 procedure | 27 (27.3) |
|  | Infection | 15 (15.2) |
|  | Bleeding | 3 (3.0) |
|  | Cerebral engagement | 1 (1.0) |
| Permanent CSF diversion, n (%) |  | 11 (11.1) |

\*CSF: Cerebrospinal Fluid; †EVD: External Ventricular Drain

Mean duration of ICU stay was 14.3 days (SD 9.2). 7 patients (3.1%) died during ICU or after discharge during the follow-up period. Most patients (91%) had a good functional outcome.

### Measure of Agreement for Radiological Scores

The agreement for the modified Fisher score was modest with a coefficient of 0.25 on the Cohen’s Kappa test [95% CI, 0.05-0.45], whereas temporal horn dilation had a higher coefficient of 0.83 [95% CI, 0.77-0.90]. Despite its good predictive value in the univariate analysis (Supplemental Material, Table S1), the modified Fisher score was not used in the multivariate model due to this low agreement.

### Predictive Values of Clinical and Radiological Variables

The different discriminating values of the potential predictors of CSF diversion are presented in Supplemental Material, Table S2. The variables we selected were the lowest GCS in the first 24 hours (AUC, 0.72 [95% CI, 0.65 - 0.78]), temporal horn dilation (AUC, 0.71 [95% CI, 0.65 - 0.78]), ventricular Hijdra score (AUC, 0.76 [95% CI, 0.69 - 0.82]), and SEBES (AUC, 0.66 [95% CI, 0.58 - 0.73]), respectively reflecting neurological status, hydrocephalus, cerebral hemorrhage, and cerebral edema.

### Guideline-Based Indication of CSF Diversion

Among the 53 patients that presented a formal indication of CSF diversion in the first 24 hours (GCS < 15 and hydrocephalus with dilation of the temporal horns), 35 (66%) were diverted. Of the 92 patients who had no formal indication, 10 (10.9%) were diverted. There were 74 patients who were in the gray area of indication, of which 29 (39.2%) were diverted (Table 3). This simplest model with 2 variables had a good AUC of 0.79 (95% CI, 0.72 - 0.85).

**Table 3:** Guideline-Based Formal Indication of CSF Diversion Within the First 24 Hours After ICU Admission.

|  | Number of patients in the<br>category, n (%) | Actual CSF*<br>diversion, n (%) |
| --- | --- | --- |
| Formal indication | 53 (24.2) | 35 (66.0) |
| Gray zone of indication | 74 (33.8) | 29 (39.2) |
| No formal indication | 92 (42.0) | 10 (10.9) |

\*CSF, Cerebrospinal Fluid.

### Predictors of CSF Diversion

The final model included 219 patients, 74 of whom had a CSF diversion within the first 24 hours of ICU admission. The variables that were independently associated with CSF diversion were the delay between the onset of symptoms and the first imaging (aOR, 0.67 [95% CI, 0.45-0.99]; p = 0.018), a GCS lower than 15 (aOR, 3.17 [95% CI, 1.42- 7.10]; p = 0.005), the ventricular Hijdra score (aOR, 1.48 [95% CI, 1.19-1.85]; p < 0.001), temporal horn dilation (aOR, 1.89 [95% CI, 1.25 - 2.87]; p = 0.002), a SEBES at 2 (aOR, 4.83 [95% CI, 1.57 - 14.87]; p = 0.030). There was significant heterogeneity between centers in this model (p < 0.001). Aae and *sine materia* SAH were not associated with CSF diversion (Table 4). The model’s discriminative performance was high, with an AUC of 0.86 [95% CI, 0.81-0.90]. The calibration of the model is shown in Supplemental Material Figure S2.

**Table 4:**
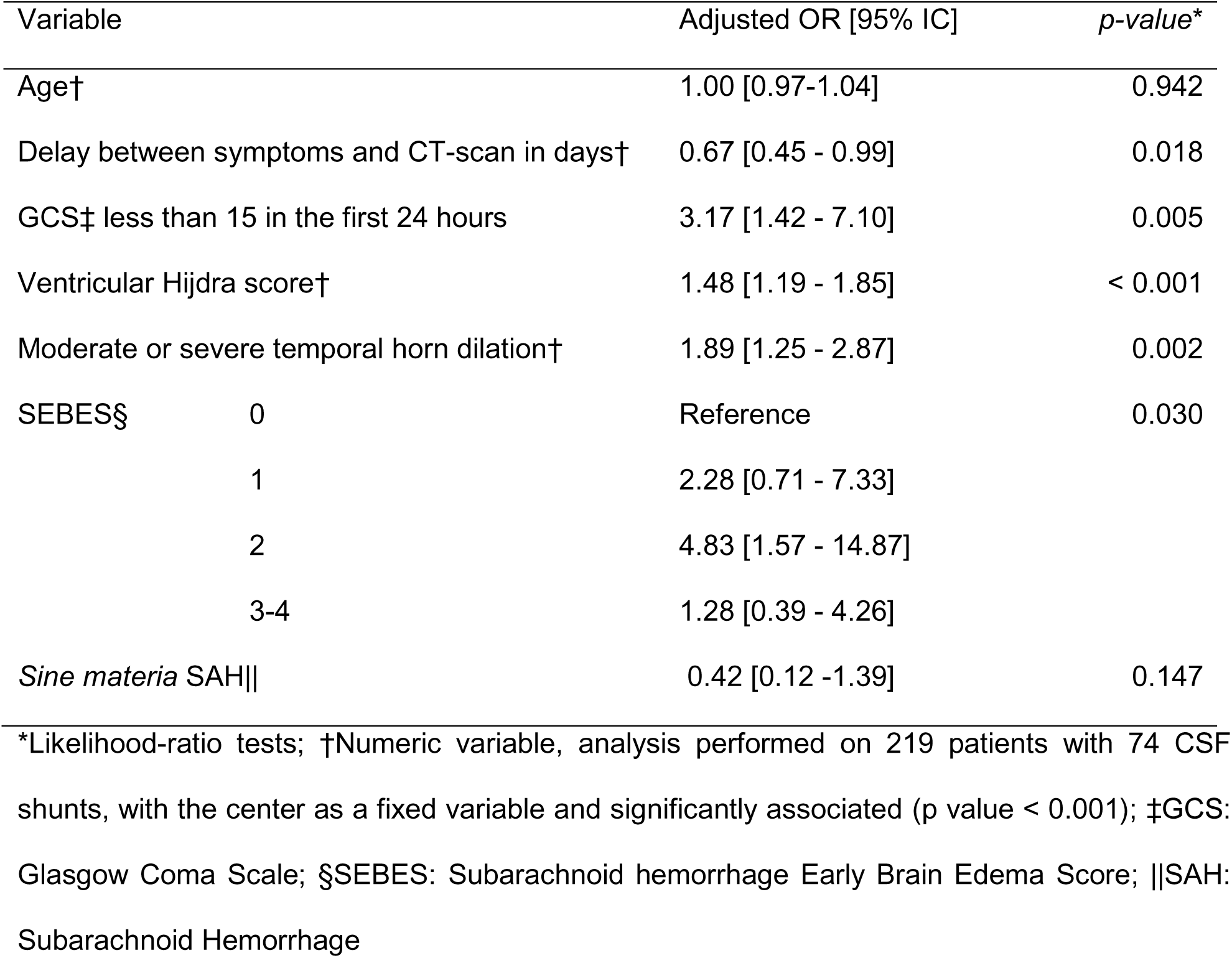
Multivariate Analysis of Predictors of CSF Diversion Within the First 24 Hours After ICU Admission.

These variables were also found to be predictors of CSF diversion during entire ICU stay (Table 5).

**Table 5:** Multivariate Analysis of Predictors of CSF Diversion During Entire ICU Stay.

| Variable | Adjusted OR [95% IC] | p-value* |
| --- | --- | --- |
| Age† | 1.03 [0.99-1.07] | 0.148 |
| Delay between symptoms and CT-scan in days† | 0.67 [0.47-0.94] | 0.008 |
| GCS‡ less than 15 in the first 24 hours | 4.00 [1.71-9.36] | 0.001 |
| Ventricular Hijdra score† | 1.76 [1.37-2.27] | < 0.001 |
| Moderate or severe temporal horn dilation† | 2.18 [1.39-3.41] | < 0.001 |
| SEBES§ |  |  |
| 0 | Reference | 0.006 |
| 1 | 4.62 [1.34-15.94] |  |
| 2 | 6.62 [1.77-24.78] |  |
| 3-4 | 2.58 [0.73-9.14] |  |
| *Sine materia* SAH\|\| | 0.58 [0.16-2.15] | 0.415 |

\*Likelihood-ratio tests; †Numeric variable, analysis performed on 219 patients with 74 CSF shunts, with the center as a fixed variable and significantly associated (p value < 0.001); ‡GCS: Glasgow Coma Scale; §SEBES: Subarachnoid hemorrhage Early Brain Edema Score; ||SAH: Subarachnoid Hemorrhage

Finally, we tested the predictive value of the PI measured by TCD at ICU admission in the final model. Due to a large amount of missing data, estimates for this model could only be made on 149 patients, and initial PI was not significantly associated with CSF diversion (aOR, 0.92 [95% CI, 0.66 - 1.28]) (Supplemental Material Table S3).

## DISCUSSION

The present study found that in clinical practice, the factors independently associated with CSF diversion in the first 24 hours following ICU admission in low-grade non traumatic SAH were: altered mental status (GCS lower than 15), hydrocephalus (temporal horn dilation), intraventricular hemorrhage (ventricular Hijdra score), the delay between the first onset of symptoms and CT-scan, and cerebral edema (SEBES). The first three of these variables (altered mental status, hydrocephalus and intraventricular hemorrhage) have been identified since the 1980s as being associated with acute hydrocephalus and therefore as risk factors of CSF diversion, as seen in international recommendations on the timing of CSF diversion^6,13,14^. However, the delay between first onset of symptoms and initial CT-scan and cerebral edema have not yet been described in known literature. A lower risk of CSF diversion with longer time from onset of symptoms and initial head imaging might be explained by a lesser severity of patients who consult later and therefore are less likely to require a CSF diversion. Cerebral edema is not described as a usual indication for CSF diversion in SAH; however, it is a cause of intracranial hypertension, of which the treatment can be CSF diversion to alleviate intracranial pressure. Since the PI measured by TCD measures the same pathophysiology as the SEBES score, it is possible that it is a predictive variable for CSF diversion, but this study was limited by the amount of missing data on this variable. There is also an important variability between centers, which can be explained by differences in local protocols and experiences due to the nonspecific wording of the international guidelines, leaving room for the clinician’s personal interpretation.

Age and *sine materia* SAH were not independently associated with CSF diversion in this study. Age was first described by Graff-Radford et al. in 1989 as being a risk factor for hydrocephalus following aneurysmal SAH, but subsequent studies on risk factors surrounding hydrocephalus have focused mainly on the risk of shunt-dependency, of which older age is indeed a risk factor, but not on acute hydrocephalus itself^3,15^. The original studies describing age as a risk factor may not have considered the natural enlargement of ventricles with age-related cerebral atrophy on head imaging, and some of the patients were diagnosed with “clinical hydrocephalus” without more information on the radiological findings. More recent studies need to be conducted to verify the risk factors specifically associated with acute hydrocephalus in the context of aneurysmal SAH.

As for *sine materia* SAH, it is classically described as having a milder clinical presentation with better short- and long-term prognosis. Given this knowledge, it would have been expected to find this variable as being associated with a lesser risk of acute hydrocephalus. However, the physiopathology of *sine materia* SAH remains unclear, with multiple hypotheses of the etiology of angiogram-negative SAH including undetected microaneurysms, bleeding from a deep vein (such as the basal vein of Rosenthal), basilar dissection, or vasospasm preventing the filling of an aneurysm^16^. There is reason to believe that the outcome of patients with *sine materia* SAH depends also on the specific presentation, with differences between perimesencephalic angiogram-negative SAH and diffuse angiogram-negative SAH, as the etiology of bleeding probably differs between these groups^17^. The physiopathology of *sine materia* and aneurysmal SAH may therefore not be as different as previously believed, which may explain that this variable is not independently associated with CSF diversion in our study.

This study shows that there is a gray area in which patients who do not have all the criteria of international recommendations (acute hydrocephalus associated with altered mental status) may have a CSF diversion, as well as patients presenting formal indication that do not have a CSF diversion, and conversely patients who present no indication who are diverted. Recent studies on the potential benefits of placing an EVD or lumbar drainage without acute hydrocephalus to reduce delayed cerebral ischemia may incite physicians to make this decision, which may in part explain that some patients without formal indication had a CSF diversion^18–20^. This further reinforces our hypothesis that international recommendations do not encapsulate all the indications of CSF diversion in patients with low-grade SAH.

This study presents strengths, including most importantly its multicentric design, allowing the study of robust and common variables across centers. This is also the first study that looks at the variables that lead to a CSF diversion in clinical practice, using radiological scores to precisely define hydrocephalus, intraventricular and subarachnoid hemorrhage, and cerebral edema. Moreover, this study analyzed over 200 patients admitted in a short period of time, limiting possible changes in local protocols over time. Finally, our models have a high discriminating quality.

The main limitation of this study is its retrospective design, which was nevertheless based on a prospective collection of data, limiting its memorization bias. Another limit is the inability to use the modified Fisher’s score, despite it being one of the most used in routine practice to identify the severity of intracranial hemorrhage. The low agreement between the score that was extracted from the Neurobase® and the score given by a neurosurgeon who was blinded to the outcome of the patient, may be explained by multiple factors, including the persisting confusion between the use of the original or the modified Fisher’s score, and the bias that the practitioners may experience collecting this data whilst knowing the patient’s outcome. This study also does not determine whether the decision to place a CSF shunt alters positively or negatively the outcome of the patients in the gray zone of indication, which needs to be further studied to establish whether a CSF diversion is beneficial, as it should be kept in mind that this invasive procedure is not without its risks.

## CONCLUSION

Altered mental status, hydrocephalus, intraventricular hemorrhage, short delay between symptoms and imaging, and cerebral edema, present in the first 24 hours of admission, are independent risk factors for CSF diversion in low-grade aneurysmal or *sine materia* SAH. External validation studies are necessary to confirm these results. The variability between centers shows that the indications for CSF diversion are not entirely established, and patients who are in a gray area of indication need to be further studied to verify the potential benefit of a CSF diversion.

## Data Availability

The data that support the findings of this study are available from the corresponding author, Marie Lejeune MD, upon reasonable request.

## NON-STANDARD ABBREVIATIONS AND ACRONYMS

SAH: Subarachnoid hemorrhage
CSF: Cerebrospinal fluid
EVD: External ventricular drain
WFNS: World Federation of Neurological Surgeons
ICU: Intensive care unit
GCS: Glasgow Coma Scale
SEBES: Subarachnoid hemorrhage Early Brain Edema Score
TCD: Transcranial Doppler
PI: Pulsatility index
mRS: modified Rankin scale
GOS: Glasgow Outcome Scale

## REFERENCES

1. Murray CJL, GBD 2021 Collaborators. Findings from the Global Burden of Disease Study 2021. Lancet 2024;403:2259–2262.

2. Steiner T, Juvela S, Unterberg A, Jung C, Forsting M, Rinkel G, et al. European Stroke Organization guidelines for the management of intracranial aneurysms and subarachnoid haemorrhage. Cerebrovasc Dis 2013;35:93–112.

3. Hoh BL, Ko NU, Amin-Hanjani S, Chou SH-Y, Cruz-Flores S, Dangayach NS, et al. 2023 Guideline for the Management of Patients With Aneurysmal Subarachnoid Hemorrhage: A Guideline From the American Heart Association/American Stroke Association. Stroke 2023;54:e314–e370.

4. Mazard T, Ritzenthaler T, Dailler F. Pravastatin may improve neurological outcome following low-grade aneurysmal subarachnoid hemorrhage. J Clin Neurosci 2022;98:11–14.

5. Mouchtouris N, Lang MJ, Barkley K, Barros G, Turpin J, Sweid A, et al. Predictors of hospital-associated complications prolonging ICU stay in patients with low-grade aneurysmal subarachnoid hemorrhage. J Neurosurg 2020;132:1829–1835.

6. Missori P, Paolini S, Peschillo S, Mancarella C, Scafa AK, Rastelli E, et al. Temporal Horn Enlargements Predict Secondary Hydrocephalus Diagnosis Earlier than Evans’ Index. Tomography 2022;8:1429–1436.

7. Mahto N, Owodunni OP, Okakpu U, Kazim SF, Varela S, Varela Y, et al. Postprocedural Complications of External Ventricular Drains: A Meta-Analysis Evaluating the Absolute Risk of Hemorrhages, Infections, and Revisions. World Neurosurg 2023;171:41–64.

8. Knol DS, van Gijn J, Kruitwagen CLJJ, Rinkel GJE. Size of third and fourth ventricle in obstructive and communicating acute hydrocephalus after aneurysmal subarachnoid hemorrhage. J Neurol 2011;258:44–49.

9. Dupont S, Rabinstein AA. CT evaluation of lateral ventricular dilatation after subarachnoid hemorrhage: baseline bicaudate index values [correction of balues]. Neurol Res 2013;35:103–106.

10. Morgan TC, Dawson J, Spengler D, Lees KR, Aldrich C, Mishra NK, et al. The Modified Graeb Score: an enhanced tool for intraventricular hemorrhage measurement and prediction of functional outcome. Stroke 2013;44:635–641.

11. Hijdra A, Brouwers PJ, Vermeulen M, van Gijn J. Grading the amount of blood on computed tomograms after subarachnoid hemorrhage. Stroke 1990;21:1156–1161.

12. Said M, Gümüs M, Herten A, Dinger TF, Chihi M, Darkwah Oppong M, et al. Subarachnoid Hemorrhage Early Brain Edema Score (SEBES) as a radiographic marker of clinically relevant intracranial hypertension and unfavorable outcome after subarachnoid hemorrhage. Eur J Neurol 2021;28:4051–4059.

13. Galera R, Greitz T. Hydrocephalus in the adult secondary to the rupture of intracranial arterial aneurysms. J Neurosurg 1970;32:634–641.

14. van Gijn J, Hijdra A, Wijdicks EF, Vermeulen M, van Crevel H. Acute hydrocephalus after aneurysmal subarachnoid hemorrhage. J Neurosurg 1985;63:355–362.

15. Graff-Radford NR, Torner J, Adams HP, Kassell NF. Factors associated with hydrocephalus after subarachnoid hemorrhage. A report of the Cooperative Aneurysm Study. Arch Neurol 1989;46:744–752.

16. Boswell S, Thorell W, Gogela S, Lyden E, Surdell D. Angiogram-negative subarachnoid hemorrhage: outcomes data and review of the literature. J Stroke Cerebrovasc Dis 2013;22:750–757.

17. Lu G-D, Wang C, Zhao L-B, Shi H-B, Liu S. Clinical Outcomes of Diffuse Angiogram-Negative Subarachnoid Hemorrhage Versus Aneurysmal Subarachnoid Hemorrhage: A Propensity Score-Matched Analysis. J Am Heart Assoc 2024;13:e031066.

18. Wolf S, Mielke D, Barner C, Malinova V, Kerz T, Wostrack M, et al. Effectiveness of Lumbar Cerebrospinal Fluid Drain Among Patients With Aneurysmal Subarachnoid Hemorrhage: A Randomized Clinical Trial. JAMA Neurol 10.1001/jamaneurol.2023.1792.

19. Al-Tamimi YZ, Bhargava D, Feltbower RG, Hall G, Goddard AJP, Quinn AC, et al. Lumbar drainage of cerebrospinal fluid after aneurysmal subarachnoid hemorrhage: a prospective, randomized, controlled trial (LUMAS). Stroke 2012;43:677–682.

20. Hulou MM, Essibayi MA, Benet A, Lawton MT. Lumbar Drainage After Aneurysmal Subarachnoid Hemorrhage: A Systematic Review and Meta-Analysis. World Neurosurg 2022;166:261–267.e9.

